# Outcomes of Mechanically Ventilated Patients with COVID-19 Associated Respiratory Failure

**DOI:** 10.1101/2020.07.16.20155580

**Authors:** Christopher S. King, Dhwani Sahjwani, A. Whitney Brown, Saad Feroz, Paula Cameron, Erik Osborn, Mehul Desai, Svetolik Djurkovic, Aditya Kasarabada, Rachel Hinerman, James Lantry, Oksana A. Shlobin, Kareem Ahmad, Vikramjit Khangoora, Shambhu Aryal, A. Claire Collins, Alan Speir, Steven Nathan

## Abstract

**Purpose:** The outcomes of patients requiring invasive mechanical ventilation for COVID-19 remain poorly defined. We sought to determine clinical characteristics and outcomes of patients with COVID-19 managed with invasive mechanical ventilation in an appropriately resourced US health care system.

**Methods:** Outcomes of COVID-19 infected patients requiring mechanical ventilation treated within the Inova Health System between March 5, 2020 and April 26, 2020 were evaluated through an electronic medical record review.

**Results:** 1023 COVID-19 positive patients were admitted to the Inova Health System during the study period. Of these, 165 (16.1%) were managed with invasive mechanical ventilation. At the time of data censoring, 63/165 patients (38.1%) had died and 102/165 (61.8%) were still alive. Of the surviving 102 patients, 17 (10.3%) remained on mechanical ventilation, 51 (30.9%) were extubated but remained hospitalized, and 34 (20.6%) had been discharged. Deceased patients were older (median age of 66 vs. 55, p <0.0001). 75.7% of patients over 70 years old had died at the time of data analysis. Conversely, % of patients age 70 or younger were still alive at the time of data analysis. Younger age, non-Caucasian race and treatment at a tertiary care center were all associated with survivor status.

**Conclusion:** Mortality of patients with COVID-19 requiring invasive mechanical ventilation is high, with particularly daunting mortality seen in patients of advanced age, even in a well-resourced health care system. A substantial proportion of patients requiring invasive mechanical ventilation were not of advanced age, and this group had a reasonable chance for recovery.

## Introduction

Since its start in late 2019 in Wuhan, China, the Coronavirus Disease 2019 (COVID-19) has blossomed into a worldwide pandemic, infecting 3 million people and killing over 200,000. (1) The rapid spread of the disease has been paralleled by an explosion of publications on the topic, with over 7,500 publications produced by a PubMed search of “COVID-19” at the start of May 2020. Despite the intense interest and effort of the medical community to better understand and treat COVID-19, substantial gaps in our knowledge of the disease remain. One particularly important area lacking clarity is the prognosis of COVID-19 patients with acute respiratory failure requiring invasive mechanical ventilation (IMV). Mortality estimates vary substantially, ranging from 16 to 97%, with multiple studies citing mortality in excess of 50%. (2-7)

These reports have led to alarming headlines in the lay press, such as “Most COVID Patients Placed On Ventilators Died, New York Study Shows,” the title of an article recently published in U.S. News and World Report. (8) However, there are significant limitations to the available literature. Much of it is derived from centers outside of the United States, where the standard of care and patient populations may differ from those seen in most United States hospitals. In addition, many of these reports come from hospitals that were experiencing a major surge in patient volumes and were forced to use suboptimal equipment and staffing models that varied considerably from typical practice. Using the data from these studies to provide counseling to families and patients with impending respiratory failure may provide an unrealistically grim estimate of the chance of survival, leading some to forego potentially life-saving treatment. We sought to delineate the survival of patients with acute respiratory failure from COVID-19 requiring IMV in a United States hospital system with a high volume of COVID-19 patients, but not surging to a capacity that outstripped the ability to provide critical care in line with the conventional standard of care.

## Methods

Data on all COVID-19 positive patients who were placed on IMV for acute respiratory failure within the Inova Health System in Northern Virginia between March 5, 2020 and April 26, 2020 was collected. Data collection was censored on May 1, 2020. The Inova Health System consists of five hospitals including a large tertiary care center and four community hospitals. COVID-19 infection was confirmed by a positive result on polymerase chain reaction testing from either a nasopharyngeal or lower respiratory tract sample. There were no transfers of COVID-19 patients into or out of the Inova Health System during the study period. Transfers within the health system to the tertiary care hospital were analyzed as a single hospitalization attributed to the accepting facility.

All data was collected from the electronic medical record (Epic®). Data collected included patient demographics (race, ethnicity age, gender), comorbidities, adjunctive respiratory treatments [paralysis, prone positioning, inhaled pulmonary vasodilators including inhaled nitric oxide, inhaled epoprostenol, and extracorporeal membrane oxygenation (ECMO)], COVID-19 targeted treatments (clinical trial enrollment, use of toculizumab, hydroxychloroquine, remdesivir, convalescent plasma), secondary infections, and outcomes [extubation, ventilator days, discharge, death, hospital length of stay, development of acute kidney injury, and need for renal replacement therapy (RRT)]. Cause of death was determined by chart review. Immunosuppressed individuals were comprised of solid organ transplant recipients, patients on active chemotherapy, and individuals on chronic immunosuppression for any other indication (at an equivalent of prednisone 20 mg daily or higher). In the event of reintubation, ventilator length of stay was calculated from date of the initial intubation until final extubation. Outcomes were unknown for a subset of patients who remained on ventilator support or hospitalized at the time of data censoring.

The initial and highest values of laboratory data including white blood cell count, ferritin, C-reactive protein (CRP), and D-dimer were also collected. Values listed as greater than or less than the maximal or minimal test value were listed as that cutoff value (e.g. d-dimer > 20 was recorded as 20). Finally, respiratory/ventilator parameters including highest positive end expiratory pressure (PEEP), highest fraction of inspired oxygen (FiO2) required and lowest ratio of pulmonary arterial oxygen tension to FiO2 (P/F ratio) were collected.

The strategy for management of acute respiratory failure was fairly homogenous across the system. Efforts were made to avoid intubation where feasible with the use of reservoir cannulas and high flow nasal cannula (HFNC). Non-invasive ventilation was largely avoided early on due to concerns about aerosolization of the virus, but was increasingly utilized over time. Inhaled nitric oxide was delivered via HFNC in a number of patients. Self-proning was incorporated where appropriate in non-intubated patients. If these measures failed and intubation was required, patients were typically managed initially with moderate PEEP (10 -12 cm H_2_0) and a lung protective ventilator strategy targeting tidal volumes of 6 mL/Kg of ideal body weight (IBW) and plateau pressures < 30 cm H_2_0. In patients with compliant lungs, tidal volumes were often liberalized to 7-8 mL/Kg IBW as long as plateau pressure remained < 30 cm H_2_0. Alternatively, some patients were switched to pressure control ventilation. Ultimately the ventilator strategy was left to the discretion of the attending intensivist. Paralysis was frequently utilized in patients with severe ARDS, defined as P/F ratio < 150. Prone positioning was also utilized in patients with a P/F ratio < 150 who required FiO2 of ≥ 60% and PEEP ≥ 10 cm H_2_0. Patients were maintained in the prone position for 16 hours or longer when performed. A conservative fluid strategy was utilized whenever possible, but was not undertaken at the expense of worsening shock. Use of inhaled pulmonary vasodilator therapy was poorly standardized and left to the discretion of the attending intensivist. The choice of sedation and analgesia was also implemented at the discretion of the attending intensivist and was targeted to a Richmond Agitation Sedation Scale (RASS) of 0 to -2. (9) Patients were considered for venovenous (VV) ECMO if age < 60 years old, on IMV < 10 days, had a P/F ratio < 100 and/or failed lung protective ventilation, despite neuromuscular blockade and prone positioning, or had recalcitrant hypercapnic acidosis affecting perfusion.

Treatments targeting COVID-19 disease were administered at the discretion of the attending physician and were subject to availability. Treatments utilized included hydroxychloroquine, toculizumab, convalescent plasma, remdesivir (either compassionate use or via clinical trial), and sarilumab via clinical trial. Need for and duration of antimicrobial agents was dictated by the attending intensivist, often with input from an Infectious Disease specialist. Use of corticosteroids and anticoagulants was poorly standardized.

This study was approved by the institutional review board (IRB # U20-05-4061). Continuous and categorical variables were presented as the median (IQR) and n (%), respectively, with the exception of length of stay data which was presented as the mean value. The Mann-Whitney U test, Chi-squared, or Fischer’s exact test were used to compare differences between survivors and non-survivors where appropriate. A p value < 0.05 was considered statistically significant. Univariate and multivariate logistic regression analysis of factors potentially associated with mortality were performed. Variables were dropped from the model through use of the likelihood ratio test. All statistical analyses were performed using STATA version 12 (StataCorp LP; College Station, TX, USA).

## Results

A total of 1023 COVID-19 positive patients were admitted in our health system during the study period. Exact numbers of patients admitted to ICU beds could not be discerned, as our health system adapted to a contingency status where critically ill patients were managed in both ICU and intermediate care beds. A total of 165 COVID-19 positive patients in our health system required invasive mechanical ventilation during the study period, representing 16.1% of admitted COVID-19 patients. At the time the data was censored, 63 patients (38.2%) had died. Table 1 describes the baseline demographics of the IMV patients. The only statistically significant demographic difference between the deceased and survivor groups (defined as alive at the time of data censoring) was age, with median ages of 66 vs. 55, respectively. Table 1 provides laboratory and ventilator data on the cohort. The only observed laboratory difference between deceased and survivor groups was a higher initial D-dimer at 2.2 versus 1.3 ng/mL (p=0.03), respectively. No significant difference was found in peak d-dimer or initial or peak CRP, ferritin and WBC. The entire cohort had severe hypoxemic failure with 52.7% having a PaO2/Fio2 ratio < 100 and 86% with a PaO2/FiO2 ratio <200. The deceased cohort had a lower nadir PaO2/FiO2 ratio at 85.7 compared to 107.7 (p=0.01) for the survivor cohort.

**Table 1.** Baseline Demographics of COVID-19 Patients Managed with Invasive Mechanical Ventilation

|  | Total<br>(n=165) | Deceased<br>(n=63, 38.2%) | Survivor*<br>(n=102, 61.8%) | p value |
| --- | --- | --- | --- | --- |
| Sex (%) | 108 male<br>(65.5%) | 39 male<br>(61.9%) | 69 male<br>(67.6%) | 0.34 |
| Age, median(IQR),<br>[range], y | 58 (50-68),<br>[23-91] | 66 (55-78),<br>[29-90] | 55 (49-64),<br>[23-91] | < 0.00001 |
| Race | White 58 (35.2%)<br>Black 39 (23.6%)<br>Asian 23 (13.9%)<br>Other 43 (26.1%)<br>No response 2<br>(1.2%) | White 39 (61.9%)<br>Black 16 (25.4%)<br>Asian 5 (7.9%)<br>Other 3 (4.8%) | White 19 (18.6%)<br>Black 23 (22.5%)<br>Asian 18 (17.6%)<br>Other 40 (39.2%)<br>No response 2<br>(1.96%) | < 0.0001^ |
| Ethnicity | Hispanic<br>59 (35.7%)<br>Non-Hispanic<br>106 (64.3%) | Hispanic<br>16 (25.4%)<br>Non-Hispanic<br>47 (74.6%) | Hispanic<br>43 (42.2%)<br>Non-Hispanic<br>59 (57.8%) | 0.03 |
| BMI, median (IQR) | 30<br>(26.1 -36.7) | 31.3<br>(25.5-37.8) | 30<br>(26.4-35.9) | 0.68 |
| Comorbidities (%) |  |  |  |  |
| HTN | 86 (52.1%) | 35 (55.6%) | 51(50%) | 0.49 |
| HLD | 47 (28.5%) | 19 (30.2%) | 28 (27.5%) | 0.71 |
| DM | 57 (34.5%) | 22 (34.9%) | 35 (34.3%) | 0.94 |
| CAD | 11 (6.7%) | 6 (9.5%) | 5 (4.9%) | 0.25 |
| ESRD | 5 (3.0%) | 4 (6.3%) | 1 (0.98%) | 0.051 |
| Immunosuppressed | 6 (3.6%) | 2 (3.2%) | 4 (3.9%) | 0.80 |
| Obesity (BMI ≥ 30) | 85 (51.5%) | 33 (52.4%) | 52 (51.0%) | 0.86 |
| Morbid Obesity<br>(BMI ≥ 35) | 49 (29.7%) | 20 (31.7%) | 29 (28.4%) | 0.65 |
| WBC (X 10<sup>9</sup>/L) |  |  |  |  |
| Initial | 7.4 (5.5-9.3) | 7.9 (5.3 -9.9) | 7.4 (5.7-9.2) | 0.85 |
| Peak | 16.4(12.5-20.7) | 17.7 (12.5-24.8) | 15.7(12.7-20.1) | 0.15 |
| CRP (mg/dL) |  |  |  |  |
| Initial | 14 (7.8-22.1) | 13.1 (7-21.1) | 15.2 (8.7-23.2) | 0.38 |
| Peak | 27.1 (17.7-33.6) | 26.5 (16.3-33.3) | 27.4 (20.1-34.1) | 0.58 |
| D-Dimer (ng/mL) |  |  |  |  |
| Initial | 1.51 (0.75-3.1) | 2.2 (1-4.2) | 1.3 (0.7-2.4) | 0.03 |
| Peak | 6.7 (3.1-13.7) | 6.5 (3-15.5) | 6.9 (2.9-13.1) | 0.76 |
| Ferritin (ng/mL) |  |  |  |  |
| Initial | 1036 (382-2151) | 1123 (444-2166) | 1033 (361-2141) | 0.94 |
| Peak | 2430 (516-3788) | 3024 (395-4770) | 2162 (615-3664) | 0.49 |
| Lowest PaO<sub>2</sub>/FiO<sub>2</sub>ratio | 97.3 (69.9-158) | 85.7 (64.2-129.7) | 107.7 (80.6-165) | 0.01 |
| ≤ 100 | n=87, 52.7% | n=40, 61.5% | n=47, 46.1% |  |
| 101-200 | n=55, 33.3% | n=15, 23.1% | n=40, 39.2% |  |
| 201-300 | n=13, 7.9% | n=3, 4.6% | n=10, 9.8% |  |
| >300 | n=10, 6.1% | n=5, 7.7% | n=5, 4.9% |  |
| Maximum PEEP | 12 (10-15) | 14 (10-16) | 12 (10-14.3) | 0.21 |

Abbreviations: BMI = Body mass index; IQR = Interquartile Range; HTN = Hypertension; HLD = Hyperlipidemia; DM = Diabetes mellitus; CAD = Coronary Artery Disease; CKD = Chronic kidney Disease WBC = White blood cell count, CRP= C-reactive protein, PEEP = Positive end-expiratory pressure.
\*Survivors were patients alive at the time of the analysis. Data are median (IQR), n (%), or n/N(%). P values calculated by Mann-Whitney U test, X<sup>2</sup> test, or Fischer's exact test, as appropriate. ^Chi squared comparing Whites versus other ethnicities.

At the time of data analysis, 63 (38.2%) of the total cohort had died. Of the remaining 102 patients, 85 (83.3%) had been successfully extubated and 34 (33.3%) of the survivors had been discharged. The mean length of stay for deceased patients was 10.8 days. The mean length of stay for patients discharged alive was 16.5 days (± 6.3 SD) (Range: 7-29 days). The average time from admission to intubation was 2.6 days (± 3.0 SD) (Range: 0-18 days); however, 43 patients (26%) were intubated on the day of admission. There was no significant difference in the mean time to intubation between the deceased patients and survivors (2.5 vs. 2.6 days, p = 0.86). The mean duration of ventilator support for extubated patients was 10.5 days (± 7.2 SD) (Range: 1-32 days). For those who died, the cause of death was hypoxemic respiratory failure in the majority of patients (n=49, 77.8%). Other causes of death included shock (n=9, 14.2%), cerebrovascular accident (n=2, 3.2%), bowel ischemia (n=1, 1.6%), subarachnoid hemorrhage (n=1, 1.6%), and complications of ECMO cannulation (n=1, 1.6%). Acute kidney injury occurred more frequently in deceased patients (n=49 (77.8%) vs. n=58 (56.9%), p=0.006).

A total of 16 patients in this cohort were treated with both ECMO and IMV, representing 9.7% of the total cohort. At the time of data analysis, 15 patients (93.75% of ECMO patients) were alive. If ECMO patients are excluded from the outcome analysis, mortality at the time of data censoring increases to 41.6% (62 of 149 patients). Table 2 summarizes the outcomes for included patients.

**Table 2.** Outcomes At the Time of Data Censoring

|  | Deceased | Survivor* | Alive, on IMV | Extubated, Hospitalized | Discharged |
| --- | --- | --- | --- | --- | --- |
| **IMV +/- ECMO (n=165)** | 63 (38.2%) | 102 (61.8%) | 17 (10.3%) | 51 (30.9%) | 34 (20.6%) |
| **IMV only (n=149)** | 62 (41.6%) | 87 (58.4%) | 14 (9.4%) | 41 (27.5%) | 32 (21.4%) |
| **ECMO only (n=16)** | 1 (6.25%) | 15 (93.75%) | 3 (18.75%) | 10 (62.5%) | 2 (12.5%) |

Abbreviations: ECMO = Extracorporeal membrane oxygenation; IMV = Invasive mechanical ventilation
\*Survivor were patients alive at the time of the analysis.

Table 3 displays the age distribution for deceased patients and survivors. Patients over 70 accounted for nearly half of the deaths in the cohort. In fact, over 75% of patients over age 70 died. In the multivariate analysis, the odds ratio of death was 1.06 for age meaning that for every one point increase in age, there was a six percent increase in the odds of death. No differences were seen with regard to gender, assessed comorbidities, or BMI. White race was found to be associated with deceased status when compared to other races. A substantial portion of the overall cohort reported Hispanic ethnicity (35.7%); Hispanic ethnicity appeared to be associated with high likelihood of survivor status in comparison to non-Hispanic ethnicity, although that association did not remain after multivariate analysis, likely due to the relationship between race and ethnicity.

**Table 3.** Age Distribution of Cohort Stratified by Survivor Status

| Age | Total | Deceased | Survivors* |
| --- | --- | --- | --- |
| ≤ 40 | 14 (8.5 %) | 4 (4.8%) | 10 (9.8%) |
| 41-50 | 32 (19.4%) | 5 (7.9%) | 27 (26.5%) |
| 51-60 | 49 (29.7%) | 17 (27.0%) | 32 (31.4%) |
| 61-70 | 37 (22.4%) | 12 (19.0%) | 25 (24.5%) |
| > 70 | 33 (20%) | 25 (39.7%) | 8 (7.8%) |

\*Survivors were patients alive at the time of the analysis

Fifty-six (54.9%) of the survivors were managed at a tertiary care center, while only 23 (36.5%) deceased patients were cared for at the same tertiary care center. Patients managed at a tertiary care center were statistically more likely to be survivors (p=0.02). Patients managed at the tertiary care center had access to both extracorporeal membrane oxygenation (ECMO) and clinical trial enrollment. Table 4 summarizes various treatments provided to the cohort of patients. Survivors were more likely to receive toculizumab or to be on ECMO, although this did not remain true after adjustment for multiple variables in the logistic regression analysis as they were correlated with access to tertiary care. Table 5 summarizes the findings of the univariate and multivariate analysis. There was a trend toward increased need for CRRT in deceased patients (33.3% versus 20.6%, p=0.07). Nearly all patients (n=152, 92.1%) received antimicrobials for some duration of time. Only 29 patients (17.6%) had confirmed, culture positive secondary infections, 11 (17.4%) in deceased patients and 18 (17.6%) in surviving patients. One patient developed *Clostridium difficle* infection.

**Table 4.** Comparison of Treatments Received by Deceased vs. Survivor Cohorts

|  | Total<br>(n=165) | Deceased<br>(n=63) | Survivors*<br>(n=102) | p value |
| --- | --- | --- | --- | --- |
| Care at Tertiary Center | 79 (47.8%) | 23 (36.5%) | 56 (54.9%) | 0.02 |
| Antimicrobials | 152 (92.1%) | 58 (92.1%) | 94 (92.2%) | 0.98 |
| Hydroxychloroquine | 132 (80%) | 45 (71.4%) | 87 (85.2%) | 0.06 |
| Tocilizumab | 48 (29.1%) | 12 (19%) | 36 (35.3%) | 0.04 |
| Convalescent Plasma | 6 (3.6%) | 1 (1.6%) | 5 (4.9%) | 0.27 |
| Clinical Trial | 20 (12.1%) | 3 (4.8%) | 17 (16.7%) | 0.32 |
| Inhaled Pulmonary Vasodilators | 69 (41.8%) | 21 (33.3%) | 48 (47%) | 0.08 |
| Paralysis | 60 (36.4%) | 21 (33.3%) | 39 (38.2%) | 0.52 |
| Prone Positioning | 100 (60.6%) | 37 (58.7%) | 63 (61.8%) | 0.70 |
| CRRT | 42 (25.5%) | 21 (33.3%) | 21 (20.6%) | 0.07 |
| ECMO | 16 (9.7%) | 1 (1.6%) | 15 (14.7%) | 0.0019 |

Abbreviations: ECMO = Extracorporeal membrane oxygenation; CRRT = Continuous renal replacement therapy \*Survivor were patients alive at the time of the analysis. Data are median (IQR), n (%), or n/N(%). P values calculated by Mann-Whitney U test, X<sup>2</sup> test, or Fischer's exact test, as appropriate.

**Table 5.**
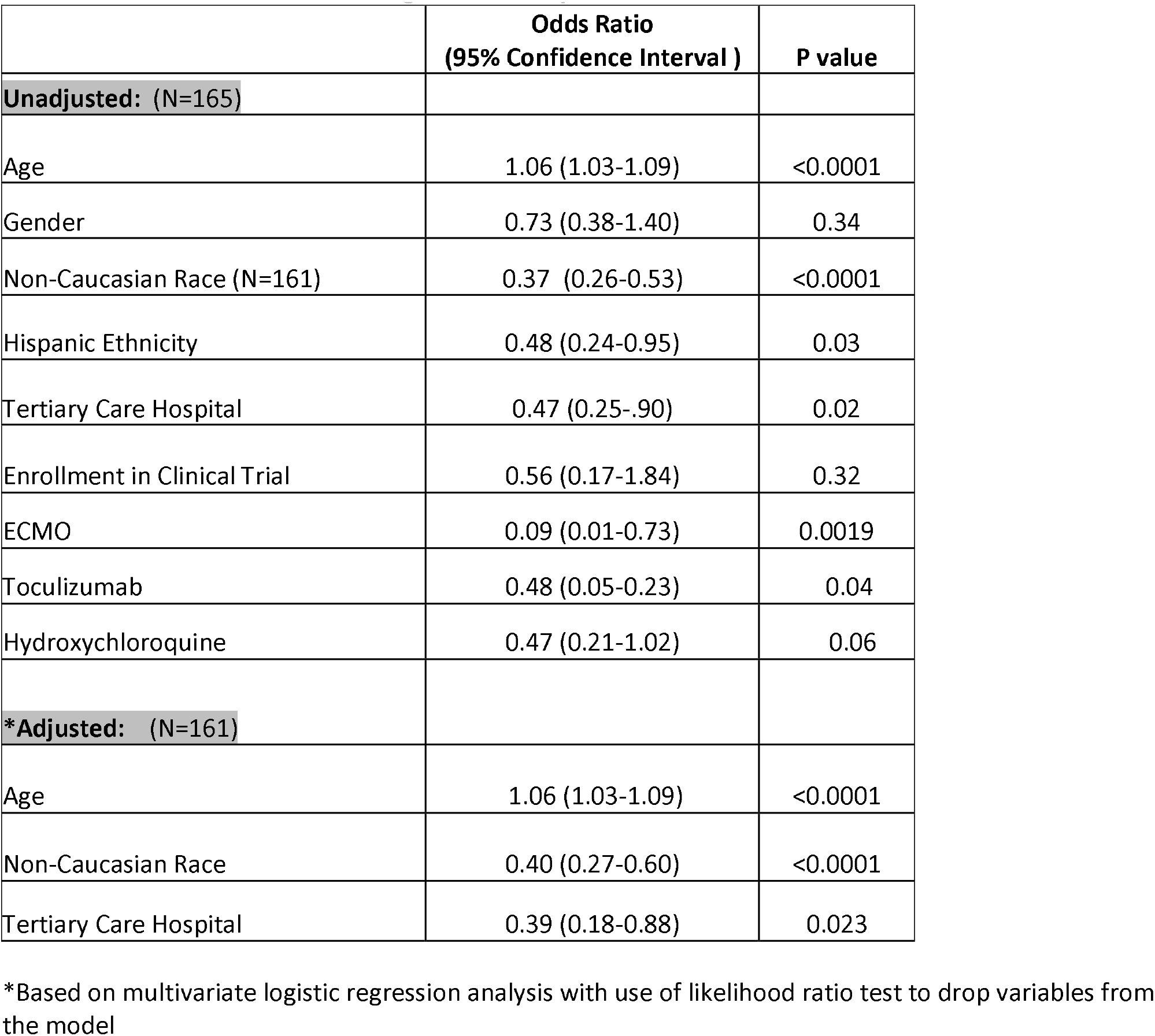
Odds Ratios of Death Among Mechanically Ventilated COVID-19 Patients

|  | **Odds Ratio**<br>**(95% Confidence Interval )** | **P value** |
| --- | --- | --- |
| **Unadjusted: (N=165)** |  |  |
| Age | 1.06 (1.03-1.09) | <0.0001 |
| Gender | 0.73 (0.38-1.40) | 0.34 |
| Non-Caucasian Race (N=161) | 0.37 (0.26-0.53) | <0.0001 |
| Hispanic Ethnicity | 0.48 (0.24-0.95) | 0.03 |
| Tertiary Care Hospital | 0.47 (0.25-.90) | 0.02 |
| Enrollment in Clinical Trial | 0.56 (0.17-1.84) | 0.32 |
| ECMO | 0.09 (0.01-0.73) | 0.0019 |
| Tocilizumab | 0.48 (0.05-0.23) | 0.04 |
| Hydroxychloroquine | 0.47 (0.21-1.02) | 0.06 |
| **\*Adjusted: (N=161)** |  |  |
| Age | 1.06 (1.03-1.09) | <0.0001 |
| Non-Caucasian Race | 0.40 (0.27-0.60) | <0.0001 |
| Tertiary Care Hospital | 0.39 (0.18-0.88) | 0.023 |

\*Based on multivariate logistic regression analysis with use of likelihood ratio test to drop variables from the model

## Discussion

The primary finding of our analysis is that mortality in COVID-19 patients requiring mechanical ventilation is high, particularly in patients of advanced age. In our cohort, the minimum mortality rate is 38.1%; however, at the time of data censoring the outcomes of a large percentage of the patients remained undetermined. The manner in which this missing data is analyzed can significantly alter the reported mortality rate. For instance, if one uses only the 97 patients with a confirmed status of death or discharge by the time of data censoring as the denominator, mortality is 64.9%. Given that an additional 30% of patients had been successfully extubated, it is likely that this is an overestimate of the true mortality rate of COVID-19 patients requiring IMV.

Despite the high mortality, the outcomes of mechanically ventilated patients in our health system compare favorably to those reported elsewhere. For instance, in the report by Richardson, et. al. on the Northwell Health System in New York, 1151 patients required IMV. At the time of their report, 24.5% of the patients had died, while only 3.3% were discharged alive, and 72% remained in the hospital. (2) If only those with a confirmed endpoint (death or discharge) from this cohort are analyzed, the reported mortality rate for patients requiring IMV is 88.1%. (2) Data from Wuhan, China reported by Zhou and colleagues found that 31 out of 32 patients (96.8%) treated with IMV died. (3) ICNARC, the Intensive Care National Audit and Research Centre from the United Kingdom, reported that 56.8% of patients treated with “advanced respiratory support”, which can include high flow oxygen, non-invasive ventilation, ECMO or IMV, died in the hospital. (7) Graselli, et. al. reported on 1591 patients hospitalized in the ICU in the Lombardy Region of Italy. (5) They do not specifically report mortality for those managed with IMV, although the majority (72%) required IMV. At the time of data reporting, their ICU mortality rate was 26%, although 58% of the patients were still in the ICU. Finally, a report from Seattle, Washington, USA included data on 24 critically ill COVID-19 patients, 18 of whom required IMV. (4) At the time of data censoring, 50% of the patients died and 5 of 18 (27.7%) were still on mechanical ventilation.

We feel it is particularly reassuring that the death rate in our cohort was not higher, given our system strategy of avoiding intubation unless patients were truly unable to maintain their oxygenation or ventilation despite aggressive management with non-invasive measures. We managed patients with self-proning, inhaled pulmonary vasodilators, and high flow oxygen to avoid intubation whenever possible. Given this approach, the cohort of patients managed with IMV was likely sicker than those reported in some of series and could be expected to have less favorable outcomes. The extreme nature of the severity of illness in our cohort is supported by the median lowest PaO2/FiO2 ratio of 97 and the fact that > 85% of the patients had a PaO2/FiO2 ratio < 200.

Patients of advanced age account for the majority of deaths in our cohort. Of 33 patients age 70 of older, 75.7% were deceased at the time the data was censored. However, only 28.8% of patients < 70 years old and 19.6% of those < age 50 were deceased at the time of data censoring. Indeed, for every increasing year of age in our cohort, there was a 6% increase in the odds of death based on our multivariate logistic regression model. This relationship between advancing age and odds of mortality is consistent with other reports. There are likely multiple reasons for this including more co-morbidities, worse baseline functional status, and variations in the aggressiveness of goals of care. A large number of patients within our cohort were offered adjunct treatments via clinical trials and/or ECMO. While the small number of patients within this cohort does not allow us to make assertions which of these treatment strategies provided the most benefit, the overall survival within our cohort likely supports the notion that availability of advanced interventions at a tertiary care center may play a significant role in improving patient outcomes.

Our study does have some limitations which should be acknowledged. First, the follow-up remains incomplete so we cannot provide conclusive outcome data. That being said, our study provides definitive outcomes data in over 60% of the cohort. We have additionally provided data on the rate of successful extubation, which has not been reported in other studies. Although ongoing follow-up would allow for definitive conclusions, we felt it was important to provide data regarding outcomes from a well-resourced, health system in a developed country. The need for additional outcomes data on IMV outcomes in COVID-19 patients was well expressed in a recent manuscript by Dr. Hannah Wunsch. (10) As she points out, prior publications with shockingly high mortality make for provocative headlines in the lay press, but may be doing a disservice to the medical community. They may invoke unjustified feelings of futility in bedside providers and prevent poorly resourced facilities from offering potentially life-saving advanced therapies. They may also prevent patients and families from accepting intensive therapies which could be potentially life-saving. Therefore, we feel our manuscript is an important addition to the available literature on outcomes of critically ill COVID-19 patients.

Another limitation of our study is that it may not be generalizable to all health systems. Our health care system has a well-resourced, well-structured and dedicated medical critical care service with high volumes and experience treating ARDS, and hence adept at the application of best practices including proning and lung protective strategies. Additionally, we have a robust and experienced, high volume ECMO program at our tertiary care hospital which bolstered our outcomes.

In conclusion, the need for IMV in COVID-19 is associated with a high mortality in patients with COVID-19. However, successful outcomes are possible, with over 70% of patients younger than 70 still alive at the time of data censoring.

## Data Availability

Data for the study is available upon request

## Acknowledgements

The study team would like to thank the physicians, nurses, respiratory therapists, pharmacists and ancillary care services who have tirelessly provide care for COVID-19 patients within the Inova health system.

